# How Strong Is a Mother’s Love? Exploring the Mediating Role of Mother-Child Relationship Quality on the Link between Child Abuse Experiences and Adult Intimate Partner Violence Perpetration

**DOI:** 10.1101/2021.08.27.21262742

**Authors:** Shuya Yin, Molly M. McLay

## Abstract

**Purpose:** Intimate partner violence (IPV) has detrimental effects on individuals’ physical and mental health. Intergenerational violence transmission theory posits that child abuse (CA) is an important determinant of later IPV perpetration due to socially learned aggression and expectations of violence in romantic relationships. However, less research exists on family protective factors like the mother-child relationship, the strength of which may mediate this link. The present study explored the association between mother-child relationships, child abuse, and adult IPV perpetration—specifically, whether mother–child relationship quality mediated the effects on the link between child abuse and later IPV.

**Methods:** This study analyzed data from individuals who completed Waves I and Wave IV of the National Longitudinal Study of Adolescent Health (N=3718 respondents). Structural equation modeling (SEM) was used to investigate mediation dynamics between CA, mother-child relationship, and adult IPV perpetration.

**Results:** Structural equation model fit the data well (CFI = .918, RMSEA = .051 [.045, .057]). The mediation model revealed that the direct effect of childhood abuse on IPV was .067 (p<.001), while the indirect effect of childhood abuse on IPV (i.e., the effect operating through the mother-child relationship) was .005 (p<.001).

**Conclusions:** While CA remains a significant risk factor for adult IPV perpetration, findings suggest that high-quality mother-child relationships may have a buffering effect and aid in preventing intergenerational violence transmission. Continued efforts to research, fund, and implement interventions that build healthy family dynamics are needed to support traumatized children into adulthood and end intergenerational violence.

## Purpose

### Background

Intimate partner violence (IPV) is a serious and prevalent public health concern. In the U.S., 1 in 4 women (27.4%) and 1 in 9 men (11.0%) have experienced sexual violence, physical violence, and/or stalking by an intimate partner in their lifetime and reported an IPV-related impact (e.g., injury, fear, concern for safety, needing services; Smith et al., 2017). These experiences can have a number of adverse effects on victim’s physical and mental health, with IPV sequelae including increased risks for functional disorders, drug abuse, sexual risk-taking behaviors, sexually transmitted diseases, depression, post-traumatic stress disorder (PTSD), and self-harming behaviors (Chrisholm et al., 2017; Gómez, 2011). IPV can even result in death; data from U.S. crime reports suggest that about 1 in 5 homicide victims are killed by an intimate partner (Centers for Disease Control and Prevention [CDC], 2014). Given these impacts, it is critically important both to examine significant risk factors for victimization and to provide early interventions to prevent IPV.

Alcohol-related problems, psychological distress, relationship satisfaction issues, and challenges within the family environment are well-documented risk factors for IPV (Caetano et al. 2001; Lipsky et al. 2005). Numerous studies have also reported that child abuse—defined as any recent act or failure to act on the part of a parent or caretaker which results in death, serious physical or psychological harm, sexual abuse, or exploitation of a child (Federal Child Abuse Prevention and Treatment Act, 2010) —is an important determinant of later IPV, both in perpetration and victimization. It is hypothesized that these risks are due to child abuse victims’ co-occurrence of behavioral problems resulting from the abuse and their acceptance of aggression as a normal part of life (Tara, Marie & Emily, 2017; Capaldi et al., 2012; Lina et al., 2013; Gómez, 2011). Child abuse has generally contributed to problematic behaviors for youth who have experienced it. These resulting behaviors—such as substance use, youth delinquency and negative emotionality—have also been shown to significantly predict later IPV (Widom, White, Czaja, & Marmorstein, 2007; Moffitt et al., 2000; Ehrensaft et al., 2003; Kim and Capaldi, 2004).

However, compared to the study of behavioral and mental health problems linking child abuse and adult IPV, only a few researchers have incorporated an exploration of parental factors in examining this association. Rakovec-Felser (2014) noted that children more easily learn aggressive behaviors through witnessing violence within nuclear families, exacerbated by chronic exposure to an abusive environment. While there has been a dearth of empirical studies in this area, several psychological and sociological theories have posited connections between child abuse and IPV within families.

### Theoretical and Empirical Explorations of Family Systems and the Child Abuse/IPV Link

Intergenerational violence transmission theory offers one explanation for the etiology of IPV within family systems (Kalmuss, 1984; Ballif-Spanvill et al., 2008; Breslin, Riggs, O’Leary, & Arias, 1990). Based on this theory, children who are exposed to family violence treat aggression as a socially learned behavior to express oneself, feel powerful, and gain control within intimate relationships in later life, thus transmitting a capacity for violence from one generation to another (Hamberger, Lohr, Bonge, & Tolin, 1997; Harned, 2001). As such, abused children may develop an expectation for violence as a normal part of romantic relationships and/or necessary to maintain control and power in their lives (Wekerle et al. 2001), opening up a pathway for IPV victimization, perpetration, or both.

Another theory that highlights the role of family as a main socializing institution and source of childhood learning is social learning theory (Bandura, 1973). Per this theory, aggression modeled between parents not only provides children with scripts for violent behaviors, but also teaches them the appropriateness and consequences of such behavior in an intimate relationship through direct and vicarious reinforcement of rewards and punishments (Bandura, 1973).

To understand and help break the cycle of intergenerational violence as posited by the aforementioned theories, family factors should be taken into consideration when studying the link between child abuse and IPV. Parent-child relationship quality may be one such factor to consider. Kaufman-Parks et al. (2018) found that positive parent-child relationships were protective against future IPV perpetration by means of a regional sample (N=950 respondents) from Ohio; individuals who reported higher parent-child relationship quality were significantly less likely to report IPV perpetration at any given point in time.

Such a notion is consistent with attachment theory (Bowlby, 1982), which argued that individuals start developing cognitive models of relationships with others based on adult caregiver-child interactions. Higher-quality parent-child relationships contribute to a safer, warmer environment for child development and may shape children’s personal cognitive models (Bowlby, 1982; Bartholomew and Horowitz, 1991). Caregiver trustworthiness and warmth can provide a foundation in infancy for youth to develop mental representations of themselves as lovable and worthy of care (Keizer, Helmerhorst & van Rijin-van, 2019). Kaufman-Parks et al. (2018) also found that the trust generated by a supportive parent–child attachment can provide youth with the confidence to build loving and secure relationships, including those with romantic partners.

Empirical evidence has additionally shown that adolescents who perceive their attachment with their parents as secure are more likely to have high levels of self-esteem (Arbona and Power, 2003; Gómez and McLaren, 2007; Wilkinson, 2004). According to Rosenberg (1965) and social bonding theory, low self-esteem weakens ties to society, decreases conformity to social norms, and may lead to delinquency, all of which are significantly predictive of IPV (Papadakaki et al., 2009). Simons et al. (1995) additionally found that—rather than the old adage that children who experience violence grow up to be violent parents—the link between childhood experiences of “harsh” parenting and later use of violence was actually mediated by parental personality (the extent to which parents displayed an antisocial orientation).

Following from this previous research exploring family factors as possible mediators, it is important to continue the exploration of how one’s family system influences the association between child abuse and adult IPV, which is imperative for uncovering intergenerational transmission mechanisms. It may be of particular interest to explore mother-child relationship quality as a potential influence on this association. Mothers are often viewed as “safe haven” attachment figures, whereas fathers are more often considered as facilitators of children’s exploration system (Dumont and Paquette 2013; Grossmann et al. 2002). As safe havens, mothers have a particular capacity to embody the warm, positive qualities outlined by Bowlby— and similar to those qualities discovered by Kaufman-Parks et al. (2018)—which may be protective against later IPV perpetration. Empirical studies of this factor, however, are few and far between.

## Present Study

Previous research has testified as to how externalized and internalized problems have mediated the link between child abuse and intimate partner violence in adulthood (Egeland, Yates, Appleyard, & Dulmen, 2002; Fergusson, Boden, & Horwood, 2005; Hussey et al., 2006; Widom, White, Czaja, & Marmorstein, 2007). Few researchers have specifically discussed the effect of family systems more broadly, and mother-child relationship quality more specifically, on IPV perpetration in adulthood. Since the literature has documented more about IPV risk factors than protective factors, increased attention is warranted on the latter, as these may provide important windows of opportunity for prevention (Capaldi et al., 2012). Such an exploration may help practitioners to design more effective preventive and psychotherapeutic interventions for the restoration of close family relationships to break the cycle of intergenerational violence transmission.

Additionally, previous research has primarily discussed the direct link between physical abuse in childhood and later IPV perpetration (Widom, Czaja & Dutton, 2014; Bevan & Higgins, 2002; Fang & Corso, 2008). Tara, Marie and Emily (2017) argued that emotional/psychological abuse is often omitted from scholarly examinations of childhood and adolescent trauma, and that this form of abuse is both highly prevalent and potentially damaging to later outcomes.

The present study specifically explored the association between mother-child relationships, child abuse, and adult IPV perpetration—specifically, whether mother–child relationship quality, as a protective factor, mediated the effects on the link between child abuse and later IPV. The present study also captured child abuse and IPV multidimensionally, correcting the omission of psychological abuse in past research. The study’s goal was to fill an empirical gap on how one particular family relationship may impact intergenerational violence transmission, both by using a nationally representative sample and robust quantitative methods.

The present study addressed following three research questions:

1. Did abused children report higher levels of IPV perpetration in adulthood?
2. Were higher quality mother-child relationships more protective against later IPV perpetration?
3. Did mother-child relationship quality mediate the link between child abuse and IPV?

## Methods

### Data and Sample

Data for this study came from the National Longitudinal Study of Adolescent Health (Add Health). Add Health is a nationally representative, school-based study of students in the United States who were in grades 7 to 12 during the 1994-1995 school year (Wave I data). Each subsequent wave combined longitudinal survey data on respondents’ social, economic, psychological, and physical well-being with contextual data on the family, neighborhood, community, school, friendships, peer groups, and romantic relationships (Harris et al., 2009). Wave IV was collected in 2008, when Wave I respondents were between the ages of 24 and 34. The current study merged publicly-available samples from Wave I and Wave IV. All interviews in these waves were conducted by audio computer-assisted self-interview technology.

Respondents were included in this study sample if they: (a) completed Wave I and Wave IV (n=5114), (b) had a sample weight (n=5114), (c) did not have missing data on covariates (n=3940), (d) were under age 18 in Wave 1 (n=3718); because child abuse was measured in Wave 1). Application of inclusion criteria resulted in a final study sample of 3718 participants.

### Measures

#### Endogenous variable

The endogenous variable of the model—IPV—was captured as a latent construct based on three observed indicator variables. The three indicators were categorical variables measured by participants’ response to three survey questions from Wave IV. Interview items were adapted from the original Conflict Tactics Scales (CTS; Straus, 1979). *Physical perpetration* referred to how often respondents slapped, hit, or kicked their partner in the past year. *Threatened perpetration* referred to how often respondents threatened partners with violence, pushed, shoved, or threw something at their partner in the past year. *Sexual perpetration* measured how often respondents insisted on unwilling sexual relations with partner in the past year. For all items, responses ranged from 0 to 7 (with 0=never; 1=happened before last year; 2=once; 3=twice; 4=3-5 times; 5=6-10 times; 6=11-20 times; 7=over 20 times).

#### Exogenous variable

The exogenous variable—child abuse— was also captured as a latent construct based on three observed indicator variables: physical abuse, sexual abuse, and psychological abuse. The measures of the three types of child abuse were developed for use in Add Health and categorized on the basis of respondents’ retrospective answers to child maltreatment before their 18^th^ birthday. *Physical abuse* indicated how often respondents experienced a parent or other adult caregiver hit them with a fist, kick them, or throw them on the floor, into a wall, or downstairs. *Sexual abuse* indicated whether respondent experienced a parent or other adult caregiver touching them in a sexual way, forcing them to touch the parent/caregiver in a sexual way, or forcing them to have sexual relations. *Psychological abuse* indicated whether respondent experienced a parent or other adult caregiver saying things that really hurt the respondent’s feelings or made them feel unwanted or unloved. For all items, responses ranged from 0 to 5 (with 0=never; 1=once; 2=twice; 3=3-5 times; 4=6-10 times; 5=over 10 times).

#### Mediating variable

*Mother-child relationship* was assessed by five items from Wave I. These items were developed for use in Add Health specifically. Mother-child relationship score was derived from the respondent’s reports of how close they felt to their mothers; if their mothers were warm and loving toward them; if they had good communication with their mothers; if they thought their mothers cared about them; and if they were satisfied with their relationships with their mothers. Answers were recorded on a five-point Likert scale from 1 (“not at all”) to 5 (“very much”). Data showing no resident mother was recorded as missing data. Cronbach’s alpha reliability for items regarding mother-child relationship was high (α = .85).

#### Control variables

Several control variables were identified for use in this study, all of which were selected due to their theoretical or empirical associations with child abuse and/or IPV (Cunradi et al., 2002; Capaldi et al., 2012; Sorenson, Upchurch, & Shen, 1996):

- Age is consistently demonstrated the protective function against IPV in adulthood (Capaldi et al., 2012). Age was defined as respondent’s age at the Wave IV interview.
- Education and income were also found to be relatively strong predictors of IPV in the United States (Cunradi et al., 2002). Education was defined as respondent’s education level at Wave IV, divided into four categories and coded 1 to 4 (1=“less than high school”; 2=“high school graduate”; 3=“some college”; 4=“college graduate”).
- Parental income was defined as respondent’s parental income as reported in Wave I. Parental income was included as categorical variable at the following levels: less than $15,000; $15,000 to $29,999; $30,000 to $69,999; and more than $70,000. (In the original dataset, 18% of observations had missing values on parental income, and these observations were excluded from this sample.)
- Sex was defined as respondent’s sex as reported in Wave I. Previous studies provided strong support for gender differences in the intergenerational transmission of violence (Fang & Corso, 2008; Houry et al., 2008; Tara et al., 2017; Li, Zhao, & Yu, 2019). Individuals of different genders may develop different problems when they faced with child abuse and IPV, such as differing levels of externalizing problems, depression, and substance use problems (White & Widom, 2003; Kratzer & Hodgins, 1997).

### Data Management and Analysis

Data management and analysis were performed by StataSE and Mplus version 7 (Muthén & Muthén, 1998–2012). To address clustering and unequal probability of selection (which could create biased estimates and false-positive hypothesis test results; see Chantala, 2006), grand sample and cluster weight variables were utilized, and these accounted for the variation in selection probability to derive population estimates in the analysis. Descriptive statistics— primarily weighted frequency, means, and standard deviations—were collected to demonstrate sample characteristics (Table 1).

**Table 1.** Descriptive Statistics of Study Population

| Variable | Percentage or Mean (SD) |
| --- | --- |
| **Exogenous Variable** |  |
| Child Abuse |  |
| Psychological | 1.50 (1.99) |
| Physical | 0.59 (1.53) |
| Sexual | 0.21 (1.05) |
| **Mediation Variable** |  |
| Mother-Child Relationship | 21.68 (4.00) |
| **Demographic Characteristics** |  |
| Age at Wave 4 | 28.69 (1.59) |
| Sex |  |
| Female | 46% |
| Male | 54% |
| Race |  |
| White | 70% |
| Black | 13% |
| Hispanic | 11% |
| Other | 6% |
| Education |  |
| Less than High School | 7% |
| High School Graduate | 22% |
| Some College | 33% |
| College Graduate | 35% |
| Parental Income |  |
| Less than \$15,000 | 16% |
| \$15,000 - \$29,999 | 22% |
| \$30,000 - \$69,999 | 46% |
| More than 70,000 | 16% |
| **Endogenous Variable** |  |
| Intimate Partner Violence (IPV) |  |
| Physical | 0.48 (1.71) |
| Threatened | 0.47 (1.71) |
| Sexual | 0.35 (1.64) |

*Note.* N=3718.

#### Analytical plan

Structural equation modeling (SEM) was employed to investigate the mediation dynamics between child abuse, mother-child relationship and IPV. SEM tests a theoretical path model against the empirical data to determine how well the model fits the actual data, as determined by model fit statistics (Schumacker & Lomax, 2010). The analysis was conducted in two steps: (1) evaluate the measurement models of child abuse and IPV; (2) analyze the direct and indirect effects of mother-child relationship on the association between child abuse and IPV in adulthood.

The first measurement model consisted of three observed indicators for child abuse and three observed variables for IPV. In order to improve model fit, we specified two pairs of correlated measurement errors: (1) physical abuse in childhood and physical perpetration in adulthood; (2) sexual abuse in childhood and sexual perpetration in childhood. In order to reduce bias in the estimation, potential covariates served as controls for the exogenous variable (child abuse) in the structural equation model. These included respondent’s education level, age in Wave IV, sex, and parental income in Wave I. Sobel’s (1986) test was utilized to approximate the standard error of an indirect effect. Good fit statistics aided in determining a final measurement model for later second-order structural equation model. Model chi-square, comparative fit index (CFI), and root mean square error of approximation (RMSEA) with a 90% confidence interval (CI) served as indicators of model fit. Good fit is typically indicated by a nonsignificant chi-square, CFI > .95, and RMSEA < .05 (upper bound of the 90% CI < .08) (Bowen & Guo, 2011).

#### Power analysis and handling of missing data

Before testing the structural equation model, power analysis was conducted to determine the minimum sample size needed to detect the effect if it exists in the full population (McHugh, 2008). The minimum sample size needed to detect an effect was calculated to be 323 respondents; the study sample far exceeded this requirement. Missing data (beyond the observations excluded due to not meeting inclusion criteria) were handled by a listwise present approach before testing the structural equation model. Data in the SEM were missing for 327 (8.7%) participants after using listwise deletion.

## Results

### Descriptive Findings

Descriptive characteristics of the sample are presented in Table 1. The average respondent was 28 years old, and the majority of them identified as white American (70%) and male (54%). The largest proportion of respondents identified their education level as college graduate (35%). Most participants’ parental income levels ranged from $30,000-$69,999 (46%). Psychological abuse was the most common form of child abuse experienced among respondents (53%), while physical IPV was the most common form of IPV perpetrated (19%).

### Model Fit Results

The final model is presented in Figure 1. Indices reflected reasonably good fit of the final model (depicted in Figure 2) with the data: CFI =0.918, RMSEA = .051 with a 90% CI [.045, .057]. This model chi-square was statistically significant (p < .001), though it is worth noting that high chi-square values based on trivial departures of the final model from the data are typical with large (n ≥ 400) samples (Bowen & Guo, 2011).

**Figure 1.**
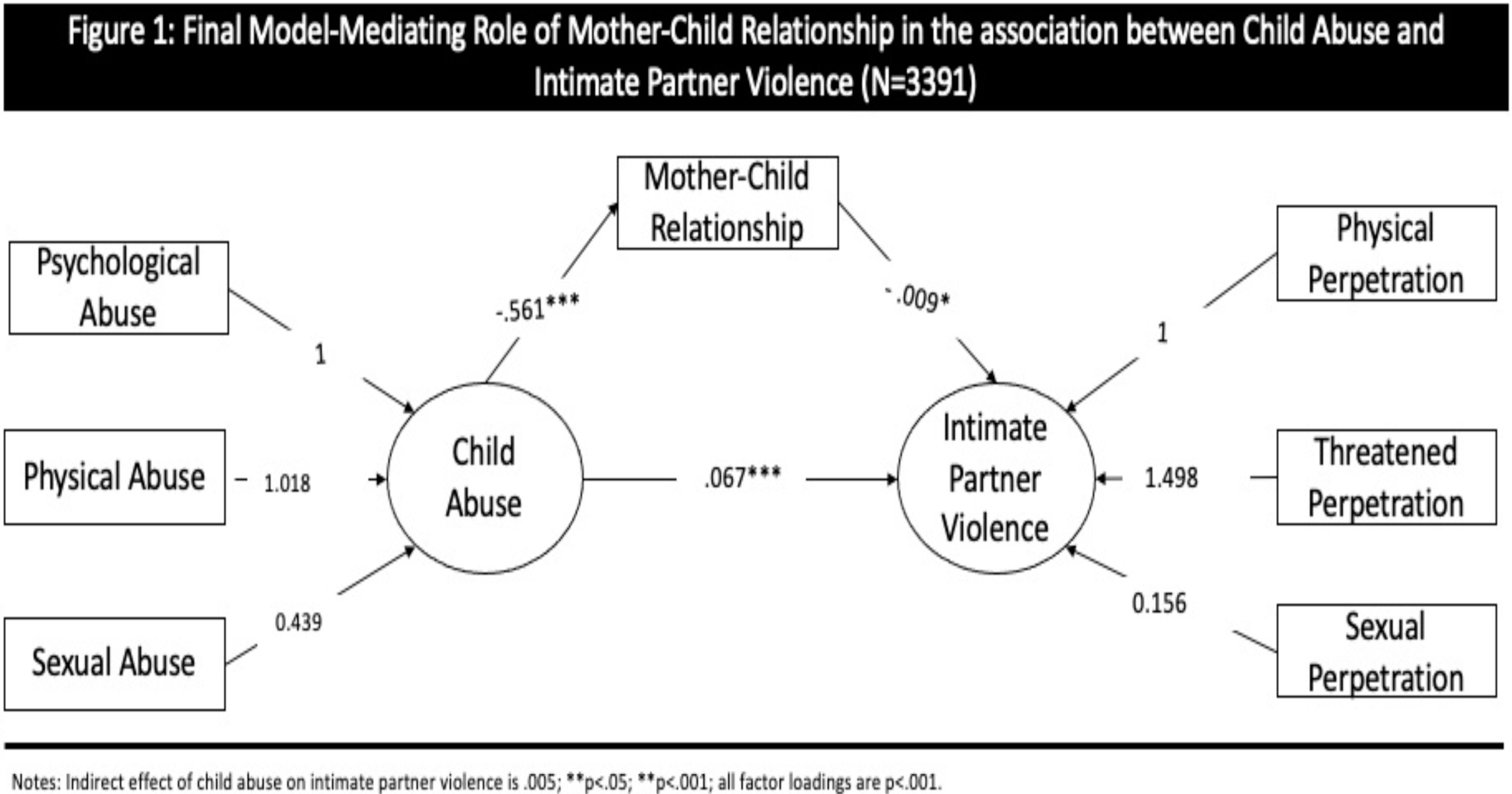
Final Model: Mediating Role of Mother-Child Relationship in the Association Between Child Abuse and Intimate Partner Violence *Note.* N=3391; indirect effect of child abuse on intimate partner violence is .005; **p<.05; ***p<.001; all factor loadings are p<.001.

### Structural Equation Modeling Results

SEM results are presented in Table 2. In the mediation model, the latent factor—child abuse—had a significant effect on both adult IPV perpetration and the mother-child relationship quality, and the former had effects that were both direct and indirect (i.e., operating through mother-child relationship quality). Other things being equal, for each additional childhood abuse experience, the quality of the mother-child relationship decreased by .561 units (p<.001), and adult IPV perpetration increased by .067 units (p<.001). In addition, every one-unit increase in the quality of the mother-child relationship decreased adult IPV perpetration by .009 units (p<.05). Each additional child abuse experience indirectly increased adult IPV perpetration through the quality of mother-child relationship by .005 units (p<.001). In other words, the mediation model revealed that the direct effect of childhood abuse on adult IPV perpetration was .067 (p<.001), while the indirect effect of childhood abuse on IPV (i.e., the effect operating through the mother-child relationship) was .005 (p<.001). Of this total effect, 93% was direct, and 6.9% was indirect.

**Table 2.** Maximum Likelihood Estimates of the Structural Equation Model Using Mother-child Relationship as Mediator

| Parameter | B | SE |
| --- | --- | --- |
| **Measurement Model** |  |  |
| Psychological Abuse → Child Abuse | 1.000 | 0.000 |
| Physical Abuse → Child Abuse | 0.489*** | 0.043 |
| Sexual Abuse → Child Abuse | 0.115*** | 0.020 |
| Physical Perpetration → Intimate Partner Violence | 1.000 | 0.000 |
| Threatened Perpetration → Intimate Partner Violence | 1.498*** | 0.159 |
| Sexual Perpetration → Intimate Partner Violence | 0.156*** | 0.044 |
| **Structural Equation Model** |  |  |
| Direct Effects |  |  |
| Child Abuse → Intimate Partner Violence | 0.067*** | 0.014 |
| Child Abuse → Mother-child Relationship | -0.561*** | 0.058 |
| Mother-Child Relationship → Intimate Partner Violence | -0.009* | 0.004 |
| Indirect Effect |  |  |
| Child Abuse → Mother-Child Relationship → Intimate Partner Violence | 0.005*** | 0.002 |
| **Model Fit Indices** |  |  |
| $\chi^2$ (df) | 233.338(24)*** | |
| CFI | 0.918 |  |
| RMSEA [90% CI] | 0.051[.045,.057] |  |

*Note.* SE = standard error; CFI = comparative fit index; RMSEA = root mean square error of approximation; CI = confidence interval. \* $p < .05$ ; \*\* $p < .01$ ; \*\*\* $p < .001$ (two-tailed test).

## Conclusions

### Summary and Interpretation of Findings

This study combined robust quantitative analysis and a nationally representative sample to explore associations between child abuse and adult IPV while controlling for individual and family factors. Study results indicated that child abuse was highly predictive of young adult IPV perpetration in this sample. These findings are consistent with intergenerational violence transmission theory, as well as with social learning theory’s explanation as to how exposure to sexual abuse and psychological abuse is associated with later IPV perpetration—because of socially learned aggression and expectations of violence in romantic relationships.

Moreover, findings indicate that the quality of the mother-child relationship is an important protective factor for preventing adult IPV perpetration, through its role as a mediator between child abuse and later IPV. While theories such as social learning theory and attachment theory have long posited the role that families play in not only exposing individuals to the use of violence, but also inculcating an acceptance and approval of the use of violence in relationships (Gelles, 1972), these results ground these suppositions in data from a large sample.

When individuals feel that parents and other caregivers love them, accept them, and are responsive to their needs, they are more likely to form secure attachments with others in later life (Bartholomew & Horowitz, 1991). Sroufe et al. (2005) characterized resilience as the development of a securely attached relationship, in that children with secure attachment to their caregiver are likely to “bounce back” from negative experiences—in the case of this study, child abuse. Children with more secure and healthier relationships with their mothers, specifically, were less likely to have engaged in IPV perpetration as adults, thus indicating a potential buffering effect of this attachment. If utilized effectively in preventive interventions, this buffer could assist in building these young people’s internal resilience in violent environments.

### Strengths and Limitations

Our research is one of the first to examine whether mother-child relationship can mediate the relationship between child abuse and IPV using national survey data. Add Health is a longitudinal, nationally representative study, which presents greater generalizability to the results and demonstrates the impact of these variables over time. It is also a very large sample, which contributes to the findings’ usefulness, as well as appropriate powering of the study. Additionally, SEM has not often been used to analyze this research area. This study has pioneered the use of this method in this research context, in sync with long-regarded—but less often empirically tested—theoretical constructs as well.

The present study was limited by data collection restraints and some missing data issues. While the sample is large and nationally representative, any retrospective measurement may contribute recall biases, and this may be the case with the study’s items around child abuse and mother-child relationship quality. Tajima et al. (2004) found that respondents tend to overestimate abuse in retrospective reports. Additionally, due to a large amount of missing data on parental income (for 18% of respondents in the original dataset), these data were excluded from the study. While information on the reasons for the data’s missingness is unknown, it is worth noting that such a large amount of missing data can present generalizability challenges and draws questions as to what important sample characteristics were not available for the complete picture of these findings. Moreover, in the present study, only mother-child relationship was measured, with no information on father-child relationship analyzed. This choice was due to the fact that variables related to father-child relationship in Add Health had over 25% missing data; the reason for this missingness is also unknown.

In order to uncover a more comprehensive picture of violence in family systems, future research could explore how paternal roles and overall parental roles function in intergenerational violence transmission. Future research could also explore how this mediation link may differ in families with only one parent, with other kinds of caregivers, or in families where parents are the same gender. Other family characteristics identified previously, such as parental marriage status and mother socioeconomic status, could be explored in future mediation studies. For example, Gelles (1976) found that poorly educated women were less likely to seek interventions after spousal abuse than were employed women with more schooling, though this sample was small (N=40). Considerations of the overall family system may be important for future research as well. Meredith et al. (1986) found that not only are individuals adversely affected by violence, but family relationships may be damaged as well, with overall family strength reduced.

Despite some limitations, this study provides important information about the complexity of child abuse and the potential protective effects of the mother-child relationship on intergenerational violence transmission. The study goes beyond long-established theories to test hypotheses with the robust, data-driven approach employed by SEM.

### Implications for Practice, Policy, and Research

In further understanding the relationship between child abuse, mother-child relationship, and later IPV, researchers and practitioners may be able to design more effective preventive and psychotherapeutic interventions to help break the cycle of intergenerational violence.

In the clinical realm, physical, sexual, and psychological abuse experienced in childhood should be paid special attention when practitioners assess clients’ previous trauma histories. This special attention paid may provide a window of opportunity to intervene on the social learning of aggression and the development of youth expectations of violence in future relationships. If children’s development of this expectation of violence in relationships can be interrupted, this could prevent future violence. Future research could explore other factors that create this expectation for violent relationships in youth as well.

At the policy level, public awareness campaigns and community, state, and system-wide violence prevention programs would do well to provide equal focus to all types of violence—not only physical and sexual, but also psychological forms of violence. These programs could also emphasize the importance of recognizing and getting support for past trauma in order to heal and prevent future trauma, both to oneself and others. Ensuring that proper resources and referrals are available as a follow-up from such programming can create additional avenues for intervention.

Implications can also be drawn from this study’s findings that parental support and involvement continue to be an important protector for abused children into adulthood. The building and/or restoration of close family relationships should be highlighted in an effort to prevent future violence. This could be done in a number of ways. Attachment-based family therapy and parent-child interaction therapy, for example, could be further promoted for use with youth and their families, and they could also modified to treat adults and their parents (Musliner & Singer, 2014). Parent-child relationship building could be utilized even earlier, in prenatal and parenting education, especially those aimed at child abuse prevention. Evidence-supported programs where parent-child connections are already a focus could be further promoted and implemented at the community, state, and federal levels.

### Final Remarks

Through the use of SEM, this study adds important empirical backing to decades of theory exploring child abuse as a significant risk factor for later IPV perpetration. While this particular confluence of theory and data may seem bleak, a glimmer of hope remains—in the form of a protective, secure mother-child relationship. When a child feels warmth, connection, and security from a mother (or potentially other caregivers), the intergenerational violence transmission cycle may in fact be stopped. If prevention programming centers this finding and works to build close family relationships between children and their loving adults, perhaps the strength of a mother’s love can stop the continuation of violence within the family system.

## Data Availability

Data is from the National Longitudinal Study of Adolescent Health (Add Health) and is publicly available.

https://addhealth.cpc.unc.edu

